# Safety and immunogenicity of the third booster dose with inactivated, viral vector, and mRNA COVID-19 vaccines in fully immunized healthy adults with inactivated vaccine

**DOI:** 10.1101/2021.12.03.21267281

**Authors:** Sitthichai Kanokudom, Suvichada Assawakosri, Nungruthai Suntronwong, Chompoonut Auphimai, Pornjarim Nilyanimit, Preeyaporn Vichaiwattana, Thanunrat Thongmee, Ritthideach Yorsaeng, Donchida Srimuan, Thaksaporn Thatsanatorn, Sirapa Klinfueng, Natthinee Sudhinaraset, Nasamon Wanlapakorn, Sittisak Honsawek, Yong Poovorawan

## Abstract

The coronavirus disease-2019 (COVID-19) pandemic has become a severe healthcare problem worldwide since the first outbreak in late December 2019. Currently, the COVID-19 vaccine has been used in many countries, but it is still unable to control the spread of severe acute respiratory syndrome coronavirus 2 (SARS-CoV-2) infection despite patients receiving full vaccination doses. Therefore, we aimed to appraise the booster effect of the different platforms of vaccines, including inactivated vaccine (BBIBP), viral vector vaccine (AZD122), and mRNA vaccine (BNT162b2) in healthy adults who received the full dose of inactivated vaccine (CoronaVac). The booster dose was safe with no serious adverse events. Moreover, the immunogenicity indicated that the booster dose with viral vector and mRNA vaccine achieved a significant proportion of Ig anti-receptor binding domain (RBD), IgG anti-RBD, and IgA anti-S1 booster response. In contrast, inactivated vaccine achieved a lower booster response than others. Consequently, the neutralization activity of vaccinated serum had a high inhibition of over 90% against SARS-CoV-2 wild-type and their variants (B.1.1.7–alpha, B.1.351–beta, and B.1.617.2–delta). In addition, IgG anti-nucleocapsid was observed only among the group that received the BBIBP booster. Our study found a significant increase in levels of interferon gamma-secreting T-cell response after the additional viral vector or mRNA booster vaccination. This study showed that administration with either viral vector (AZD1222) or mRNA (BNT162b2) boosters in individuals with a history of two doses of inactivated vaccine (CoronaVac) obtained great immunogenicity with acceptable adverse events.

## 1. Introduction

Severe acute respiratory syndrome coronavirus 2 (SARS-CoV-2) belongs to the family of coronaviruses in the genus of *Betacoronavirus*. SARS-CoV-2 is rapidly transmitted as airborne contagious droplets, when in close contact with an infected carrier. Since the beginning of the coronavirus disease 2019 (COVID-19) outbreak in Wuhan, China, in December 2019, this virus had been dramatically transmitted worldwide. The World Health Organization (WHO) declared COVID-19 a pandemic on the 11 March 2020. Globally, as of 2 December 2021, over 263 billion confirmed cases of COVID-19 have been recorded, including approximately 5.2 million deaths [1]. Several studies have been reported regarding the efficacy of the COVID-19 vaccine to help minimize disease severity and mortality [2–4]. Therefore, the COVID-19 vaccination has been administered on a global scale. Thailand had begun to inoculate most of the population with an inactivated vaccine (CoronaVac; Sinovac Life Sciences, Beijing, China) mainly since the end of February 2021, before the arrival of the viral-vector vaccine, AZD1222 (University of Oxford/AstraZeneca, Oxford, UK) from Siam bioscience (AstraZeneca [Thailand] Co., Ltd, Nonthaburi, Thailand). Later on, Thailand launched the first mass vaccination program with many vaccines during June 2021. Furthermore, inactivated vaccine, BBIBP (Beijing Institute of Biological Products Co., Ltd.; Sinopharm, Beijing, China); viral vector vaccine, Ad26.CoV2.S (Janssen Biotech Inc., PA); mRNA vaccine consisting of BNT162b2 (Pfizer-BioNTech Inc., NY); and mRNA-1273 (Moderna, Inc., MA) have been approved by the Thai Food and Drug Administration (FDA) [5].

However, breakthrough infection has been reported despite having received the full dose of CoronaVac in medical healthcare workers [6]. Although the administration of CoronaVac was achieved in Bangkok, we observed that the low vaccine efficacy affected control efforts to limit the spread of SARS-CoV-2 infection [7]. In April 2021, the alpha (B.1.1.7) variant was prominent in Thailand. The delta (B.1.617.2) variant became the most prevalent one after July 2021 [8] and also spread globally, as recognized by WHO and the Centers for Disease Control and Prevention (CDC) [9–10]. Currently, the delta variant has overtaken the original COVID-19 virus strain as the most widespread strain in the pandemic, even in regions with high vaccination [10–12]. Therefore, the booster dose of SARS-CoV-2 is urgently required to stabilize the rising numbers of infected individuals. The preliminary immunogenicity data from the real-world immunization has indicated that the booster dose of AZD1222 in the CoronaVac cohort elicited a high antibody titer and neutralization activity against SARS-CoV-2 and their variants [13]. In addition, animal model studies have shown that the booster dose with inactivated, adenoviral vector, and mRNA vaccines evoked a high titer of neutralizing antibody [14]. Therefore, in this study, we aimed to investigate the reactogenicity and immunogenicity of the third booster dose with the same and different COVID-19 platform vaccines.

## 2. Materials and Methods

### 2.1 Study design

This participant cohort study comprised three groups of healthy Thai adult participants (age: ≥18 years). A total of 177 healthy subjects who received two doses of the inactivated COVID-19 vaccine, CoronaVac (CV) (Sinovac Biotech co., LTD., Beijing, China), were divided into three groups. The first group received the inactivated vaccine BBIBP-CorV (Beijing Institute of Biological Products Co., Ltd.; Sinopharm, Beijing, China) (n=60); the second group received the viral-vector vaccine– ChAdOx1-S/nCoV-19, also called AZD1222 (University of Oxford/AstraZeneca, Oxford, UK) (n=57); and the third group received an mRNA vaccine called BNT162b2 (Pfizer-BioNTech Inc, NY) (n=60), at 3–4 months after receipt of the first dose (Figure S1). After receiving the additional dose, all study participants were monitored for safety and immunogenicity

The study protocol was approved by the Institutional Review Board (IRB), Faculty of Medicine, Chulalongkorn University (IRB number 546/64), and this trial has been registered with the Thai Clinical Trials Registry (TCTR 20210910002). Informed consent was obtained before participant enrollment. The study was conducted according to the Declaration of Helsinki and the principle of Good Clinical Practice Guidelines (ICH-GCP).

The participants were initially recruited after ensuring that there was no history of SARS-CoV-2 infection. Blood samples were collected prior to vaccination (day 0, baseline) and after receiving the booster dose (day 14 and day 28).

### 2.2 Vaccines

BBIBP-CorV (referred to as BBIBP) is an inactivated vaccine developed from whole SARS-CoV-2 stain HB02. Briefly, the HB02 strain is obtained by passaging and purification in Vero cells. Consequently, the whole virion is inactivated in β-propionolactone and further absorbed with aluminum hydroxide. One dose (0.5 mL) contains 6.50 U [15].

ChAdOx1-S/nCoV-19 (referred to as AZD1222) is a non-replicating chimpanzee adenovirus Oxford 1 vector vaccine presenting the SARS-CoV-2 spike protein (AZD1222). The virion is produced in genetically modified HEK293 cells. One dose (0.5 mL) contains 5×10^10^ infectious units [16].

BNT162b is a lipid nanoparticle containing modified RNA encoding the SARS-CoV-2 full-length spike, modified by two proline mutations to lock it in the prefusion conformation. One dose (0.3 mL) contains 30 μg [17].

### 2.3 Reactogenicity assessment

The subject was continuously monitoring for details of AEs following immunization (AEFIs) within 7 days, by self-assessment records via online or paper-based questionnaires. Explanation about data collection was given to participants by trained investigators during the initial visit, and both local, systemic, and any AEs were recorded.

### 2.4. Laboratory measurements

#### 2.4.1 Immunoglobulin and IgG anti-RBD assay

The serum fraction from the participant was analyzed for total immunoglobulin (Ig) specific to the receptor-binding domain (RBD) of the SARS-CoV-2 spike (S) protein using an electrochemiluminescence immunoassay (ECLIA) – Elecsys SARS-CoV-2 S according to the manufacturer’s instructions (Roche Diagnostics, Basel, Switzerland). The quantitative immunoglobulin IgG titer was reported as unit per milliliter (U/mL). The IgG anti-RBD was tested using chemiluminescent microparticle immunoassay (CMIA) – SARS-CoV-2 IgG II Quant assay (Abbott Laboratories, Abbott Park, Il.) according to the manufacturer’s instructions. The results are quantitative and given as arbitrary units per milliliter (AU/mL). Then, the value was multiplied by 0.142 to convert it to binding antibody units per milliliter (BAU/mL).

#### 2.4.2 IgG anti N assay

For the SARS-CoV-2 anti-nucleocapsid (anti-N) IgG, the serum fraction was also tested using the CMIA (Abbott Diagnostics, Sligo, Ireland). The semi-quantitative results are reported in the unit of sample per calibrator or index (S/C) and interpreted following the manufacturer’s instruction. For interpretation, S/C≥1.4 was defined as positive, and S/C<1.4 as negative.

#### 2.4.3. IgA anti S1 assay

The SARS-CoV2-2 anti-S1 IgA was monitored using an enzyme-linked immunosorbent assay (ELISA) (Euroimmun, Lübeck, Germany). Each kit contains microplate strips with 8 break-off reagent wells coated with recombinant structural protein of SARS-CoV-2. Briefly, in the first reaction step, diluted patient samples are incubated in the wells. In the case of positive samples, specific antibodies will bind to the antigens. A second incubation is carried out using an enzyme-conjugated antihuman IgA catalyzing a color reaction to detect the bound antibodies. The color intensity after the stop reaction was evaluated at 450 nm. The semi-quantitative analysis can be interpreted by calculating a ratio of optical density between the sample and the calibrator. The upper limit ratio (S/C)>9 was reported as 9.0.

#### 2.4.4. Neutralization assay

Serum samples were also evaluated for neutralizing activity against the SARS-CoV-2 wild-type and variant of concern (VOCs)— B.1.1.7 (alpha), B.1.351 (beta), and B.1.617.2 (delta) using an ELISA-based surrogate virus neutralization test (sVNT). Additionally, cPass SARS-CoV-2 neutralizing antibody detection kit (GenScript, Piscataway, NJ) was used for all strains. The recombinant RBD from B.1.1.7 (containing N501Y), B.1.351 (containing N501Y, E484K, and K417N), and B.1.617.2 (containing L452R and T478K) were also used with this kit. Briefly, the serum samples were diluted 1:10 with buffer and incubated with RBD conjugated to horseradish peroxidase for 30 min at 37°C. Next, 100 μL of the sample mixture was added to a capture plate pre-coated with human angiotensin-converting enzyme 2 (ACE2) and incubated for 15 min at 37°C. After washing, 100 μL of 3,3′,5,5′-tetramethylbenzidine (TMB) solution was added, and the plate was incubated in the dark for 15 min at room temperature. After the addition of 50 μL stop solution, samples were read at 450 nm. The ability of a serum to inhibit binding between RBD and ACE2 was calculated as a percentage as follows: 1 - (average OD of sample/average OD of negative control) × 100.

#### 2.4.5 SARS-CoV-2 stimulating IFN-γ assay

Heparinized whole blood was collected to the blood collection tube following the manufacturer’s instruction and incubated at 37°C for 21 h (QuantiFERON Human IFN-γ SARS-CoV-2, Qiagen). This assay consists of two sets of SARS-CoV-2 S antigens. The Ag1 tube contains CD4+ epitopes derived from the S1 subunit (RBD), the Ag2 tube contains CD4+and CD8+ epitopes from the S1 and S2 [18]. Moreover, the plasma fraction was used to determine the concentration of IFN-γ (IU/mL) using QuantiFERON® ELISA. Absorbance was measured at 450 nm and calculated using IFN-γ standard curve using the QuantiFERON R&D Analysis Software. The QFN ELISA’s detection limit is 0.065 IU/mL, while the maximum limit was 10.0 IU/mL.

### 2.5. Statistical analysis

The graphical representation and statistical analyses were carried out using GraphPad Prism version 7.0 for Microsoft Windows. Categorical analyses on age and sex were performed using the chi-square test and Welch’s ANOVA. IgG-specific RBD was designated as geometric mean titers (GMT) with a 95% confidence interval (CI). Other parameters were presented as medians with interquartile range. The difference in antibody titers, S/C, and percentage inhibition and IU/mL minus nil between groups was calculated using the Kruskal–Wallis test or Wilcoxon signed-rank test (non-parametric) with multiple comparison adjustments. A p-value <0.05 was considered to indicate statistical significance.

## 3. Results

### 3.1 Demographic data

Healthy participants aged ≥18 years who received two doses of CV were recruited by physicians or trained research nurses for additional dose with inactivated, viral vector or mRNA COVID-19 vaccine after 3-4 months from the first dose of CV. All participants had no history of underlying disease (inactive disease), immunocompromise, or any ongoing immunosuppressive therapy. The baseline demographic characteristics of all participants in the three groups, namely those who received BBIBP, AZD1222, or BNT162b2 vaccines were generally comparable (Table 1). The mean age of participants who received BBIBP, AZD1222, and BNT162b2 was 42.7, 41.6, and 44.2 years, respectively. No significant difference in age and sex was observed among the study groups. In the AZD1222 regimen, one participant was excluded as they developed SARS-CoV-2 infection, which was validated by nasopharyngeal swab and real-time polymerase chain reaction on day 7 after vaccination. This participant was asymptomatic until leaving the alternative quarantine station. Additionally, one patient was lost to follow-up during the study period, while another one who received the BNT162b2 vaccine was also lost to follow-up (Table 1).

**Table 1.** Demographics and characteristics of the vaccinated cohorts.

|  | **BBIBP** | **AZD1222** | **BNT162b2** |
| --- | --- | --- | --- |
| Total number (n) of participant | 60 | 57 | 60 |
| Mean age (year, range) | 42.7 (20–62) | 41.6 (21–59) | 44.2 (25–58) |
| Sex |  |  |  |
| Male (%) | 30/60 (50.0%) | 29/57 (50.9%) | 24/60 (40.0%) |
| Female (%) | 30/60 (50.0%) | 28/57 (49.1%) | 36/60 (60.0%) |
| Underlying disease (%) |  |  |  |
| Allergy | 5/60 (10.0%) | 2/57 (3.5%) | 4/60 (6.7%) |
| Breast cancer <sup>#</sup> | – | – | 1/60 (1.7%) |
| Cardiovascular diseases | 1/60 (1.7%) <sup>1</sup> | 2/57 (3.5%) <sup>2</sup> | – |
| Diabetes Mellitus | 1/60 (1.7%) | 2/57 (3.5%) | – |
| Dyslipidemia | 4/60 (6.7%) | 1/57 (1.8%) | 6/60 (10%) |
| Hypertension | 4/60 (6.7%) | 2/57 (3.5%) | 2/60 (3.3%) |
| Other (Gastritis, Migraine, Thyroid disease, etc.) <sup>#</sup> | 2/60 (3.3%) | 3/57 (5.2%) | 2/60 (3.3%) |
| Follow-up |  |  |  |
| Second visit (two weeks) |  |  |  |
| Mean (day, range) | 14.2 (13–18) | 14.1 (14–17) | 14.1 (14–19) |
| SARS-CoV-2 infection (n) | 0 | 1 | 1 |
| Lost to follow-up (n) | 0 | 0 | 0 |
| Third visits (four weeks) |  |  |  |
| Mean (day, range) | 28.3 (25–31) | 27.9 (27–28) | 28.1 (28–31) |
| SARS-CoV-2 infection (n) | 0 | 1 | 0 |
| Lost to follow-up (n) | 0 | 1 | 1 |

<sup>#</sup> Inactive disease – no medication involving immunosuppressant.
<sup>1</sup> Coronary artery disease; <sup>2</sup> Heart disease.

### 3.2 Reactogenicity data of the cohort receiving a different type of SARS-CoV-2 vaccine

Between March 2021 and May 2021, the Thai participants who received two doses of CV approximately 3–4 months apart were recruited for a third booster vaccine. The reactogenicity data of this cohort was significantly less with respect to solicited local and systematic adverse events (AEs) in those who received with inactivated vaccine (BBIBP) as compared with others. Moreover, most participants who received BBIBP developed minimal post-vaccination AEs (Figure 1A). The most common solicited local AE after the booster was pain at the injection site: BBIBP group (36.7% mild, 5.0% moderate AE had peaked on day 0) (Figure 1A); AZD1222 group (52.6% mild, 26.3% moderate, and 5.3% severe AE had peaked on day 1) (Figure 1B); and BNT162b2 (60.0% mild, 30.0% moderate, and 1.7% severe AE had peaked on day 1) (Figure 1C). The most common systemic AE after the booster dose was myalgia: BBIBP group (21.7% mild, 3.3% moderate AE had peaked on day 0) (Figure 1A); AZD1222 group (43.9% mild, 22.8% moderate, 5.3% severe AE had peaked on day 1) (Figure 1B); and BNT162b2 group (38.3% mild, 16.7% moderate; 1.7% severe AE had peaked on day 1) (Figure 1C). The local and systemic AEs were reduced within a few days after vaccination (Figure 1). Further analysis revealed that the AEs after taking the BBIBP vaccine were considerably lower than the other two vaccine types, except for vomiting. Joint pain was substantial in 40.4% of the total participants receiving the AZD1222 vaccine (Supplementary Table S1).

**Figure 1.**
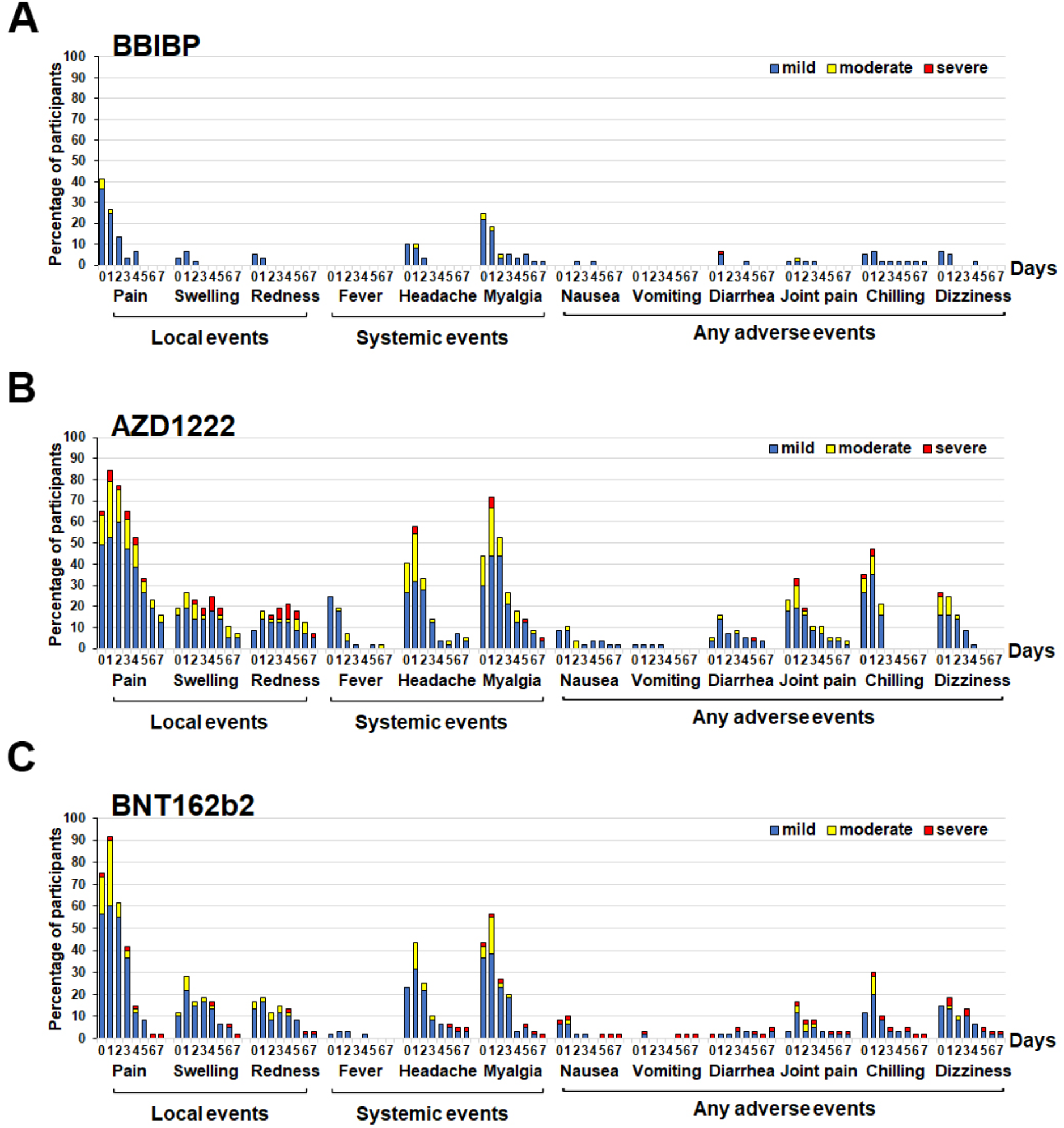
Reactogenicity of a booster dose of SARS-CoV-2 vaccines within 7 days of vaccination. A booster dose of the inactivated vaccine – BBIBP (A), viral vector vaccine – AZD1222 (B), and mRNA vaccine – BNT162b2 (C). The percentage of participants who recorded local, systemic, and any adverse events is shown on the Y-axis. Fever is defined as mild: 38.0°C to <38.5°C; moderate: 38.5°C to <39.0°C; severe: ≥39.0°C. For local and systemic symptoms, grading was classified as mild – easily tolerated with no limitation on regular activity; moderate – some limitation of daily activity; severe – unable to perform regular daily activity [18].

### 3.3 Antibody assay after booster dose with a different type of SARS-CoV-2 vaccine

To investigate the total immunoglobulin anti-RBD of SARS-CoV2, mainly IgG and IgG anti RBD were compared among all groups using the Kruskal–Wallis test with multiple comparison adjustment (Figure 2A-B, Supplementary Table S2). The geometric mean titer (GMT) of the total Ig anti-RBD and IgG anti-RBD of the three groups showed similar levels before the booster dose (baseline, day 0). The third dose with BBIBP, AZD1222, or BNT162b2 vaccines significantly elicited total Ig anti-RBD GMTs for 1,073, 9,865, or 20,787 U/mL at 14 days post vaccination, respectively (*p<*0.0001). Moreover, the immunoglobulin anti-RBD was slightly reduced at 28 days to determine the level of IgG anti-RBD. The trend of IgG anti-RBD was consistent with the immunoglobulin anti-RBD results (Figure 2A-B, Supplementary Table S2). The data indicated that the BNT162 vaccine has the highest immunization whereas BBIBP showed the lowest immunization. Compared to the antibody titer at 14 days between the AZD1222 and BNT162b recipients, we found that there were no significant differences in the GMTs of the total Ig anti-RBD and IgG anti-RBD.

**Figure 2.**
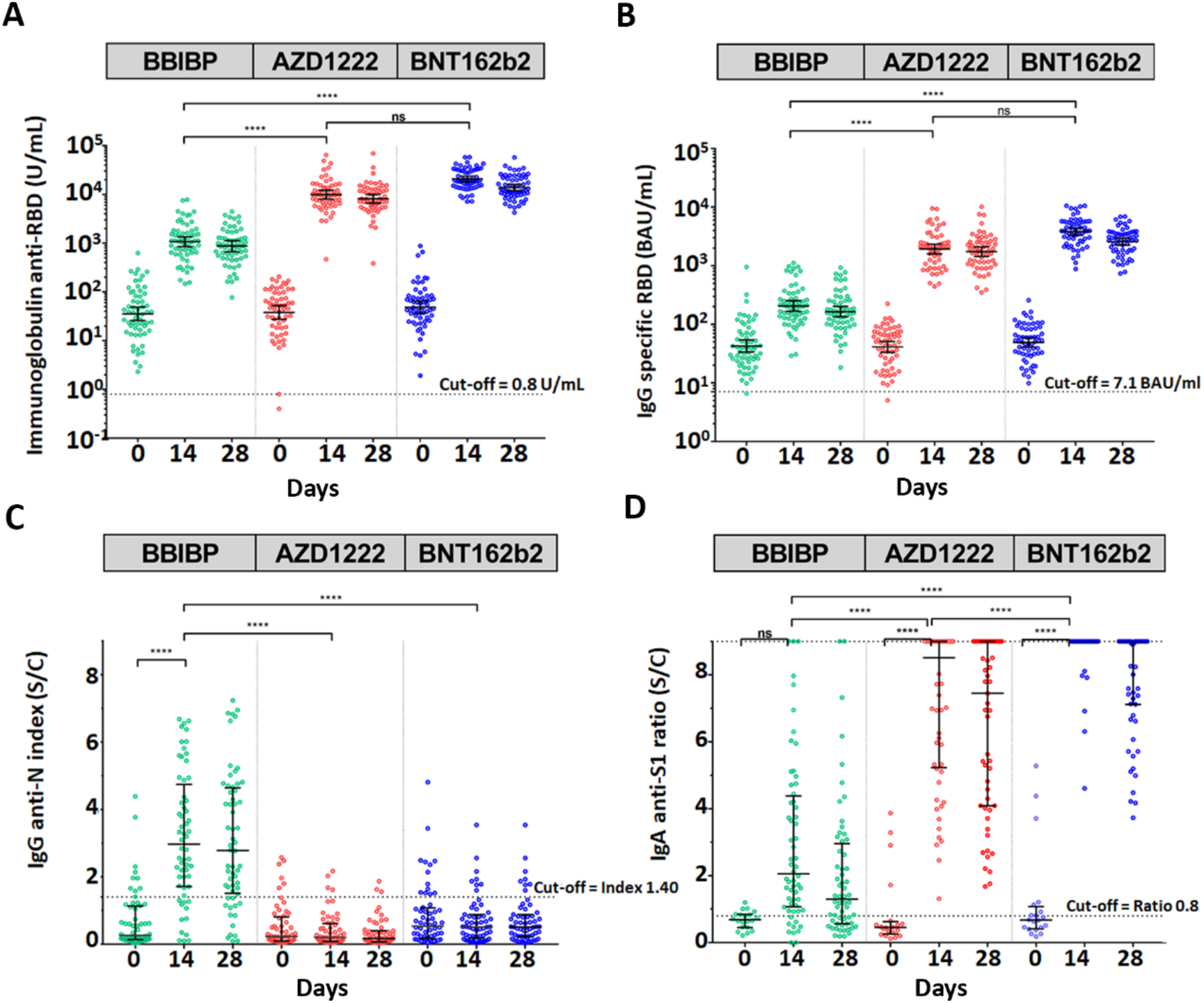
Antibody response against SARS-CoV-2 assay. The circulating total immunoglobulin anti-RBD of SARS-CoV-2 (U/mL) (A). The circulating IgG-specific RBD of SARS-CoV-2 (BAU/mL) (B). IgG anti-N of SARS-CoV-2 index (S/C) (C), IgA anti-S1 of SARS-CoV-2 ratio (S/C) (D). The serum samples were obtained from participants who received two completed doses of the inactivated vaccine, CoronaVac, followed by the inactivated vaccine, BBIBP (green), the viral-vector vaccine, AZD1222 (red), or the mRNA vaccine, BNT162b2 (blue) at 3–4 months after the first dose. Lines represent GMT (95% CI). ns indicates no statistical difference; *p*<0.0001 (****).

The median result showed that IgG anti N (nucleocapsid) was seronegative (cut-off <1.4) at baseline. The data represented that a total of 177 participants were not previously infected with SARS-CoV-2 before the booster dose. Moreover, only BBIBP could significantly elicit IgG anti N (Figure 2C, Supplementary Table S2). Besides, the median of IgG anti-N could be detected, but below to the cut-off ratio in the AZD1222 and BNT162b groups. The residual IgG anti-N was possibly obtained from the previous two doses of CoronaVac (Figure 2C).

There was no significant difference of IgA anti-S1 among the three patient groups at baseline. Interestingly, the booster dose could significantly increase IgA anti-S1 (*p<*0.0001) after receiving AZD1222 and BNT162b. Although BBIBP could elicit over 2-folds of IgA anti-S1-ratio at 14 days compared with baseline, the difference was not significant (Figure 2D, Supplementary Table S2). Additionally, the IgA anti-S1 level decayed at day 28 with Ig/IgG anti-RBD (Figure 2, Supplementary Table S2).

### 3.4 Neutralization assay against SARS-CoV-2 wild-type and variant of concerns

Surrogate virus neutralization assay was performed to determine whether the vaccinated serum has functionally inhibited the SARS-CoV-2 binding. The data of neutralization activity against SARS-CoV-2 of vaccinated serum was in accordance with the Ig RBD and IgG RBD results. The results showed that the booster with either AZD1222 or BNT162b2 had inhibition activity against wide-type (WT) and variant strains over 90% on day 28, whereas the booster with BBIBP was significantly lower in inhibition activity against SARS-CoV-2 WT and their variant strains than the two other two vaccines (Figure 3 A-C. Supplementary Table S2). There was no comparison of neutralizing activity between the AZD1222 and BNT162b groups, as the neutralization inhibition was presented at the upper limit of the assay.

**Figure 3.**
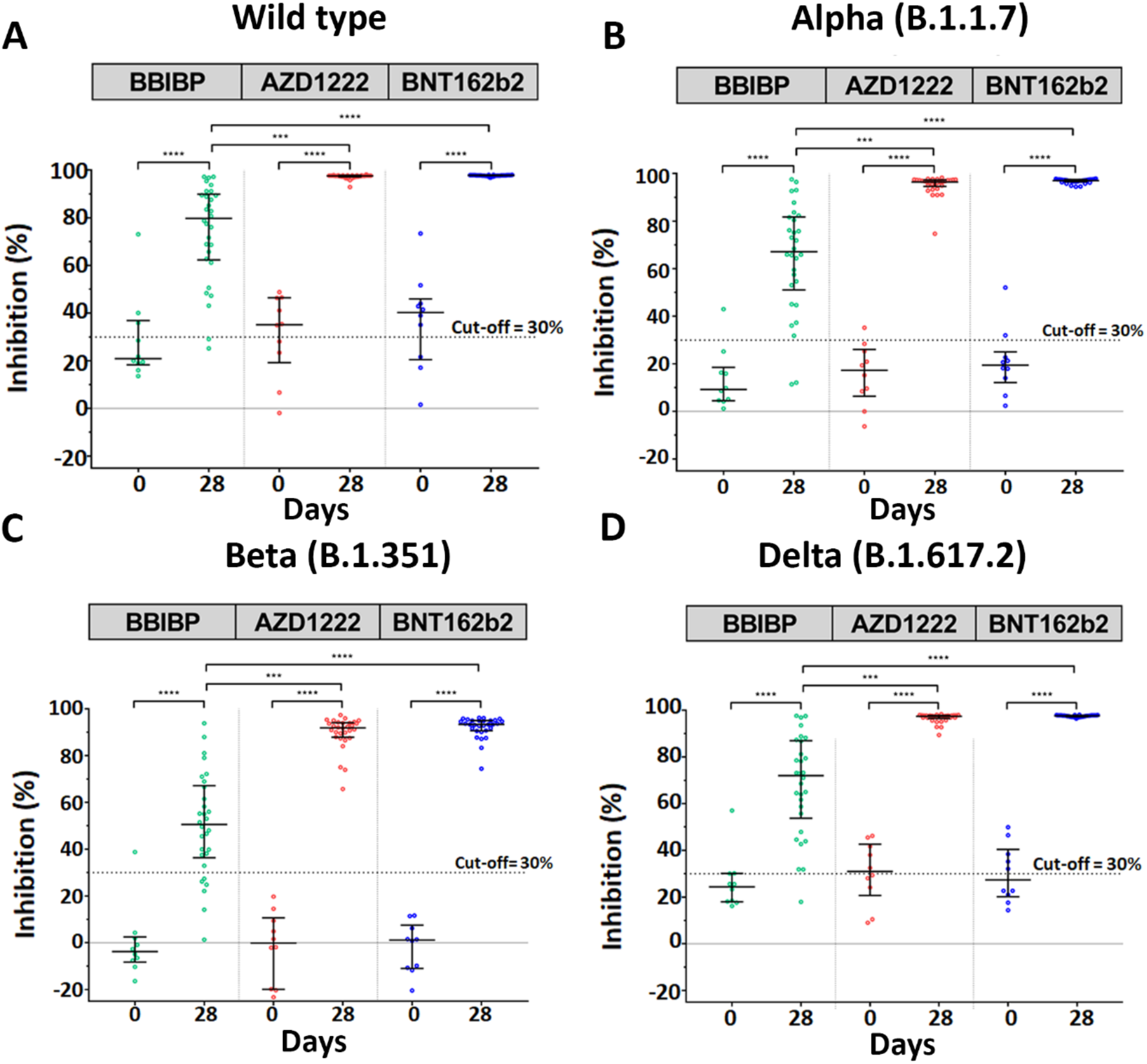
Neutralization assay by surrogate viral neutralizing test (sVNT). The serum samples were obtained from participants who received two completed doses of the inactivated vaccine, CoronaVac, followed by either the inactivated vaccine, BBIBP (green, A), the viral-vector vaccine, AZD1222 (red, B), or the mRNA vaccine, BNT162b2 (blue, C) at 3–4 months after the first dose. Lines represent median (IQR). ns indicates no significant difference; *p-*value<0.001 (***), 0.0001 (****).

According to the data, the booster with either viral vector or mRNA was more favorable given the high immunogenicity against SARS-CoV-2 WT and their variants.

### 3.5 SARS-CoV-2 IFN-γ stimulation

To address whether the vaccination generated a T-cell response, a whole blood interferon-gamma release assay (IGRA) using QuantiFERON (QFN) SARS-CoV-2-ELISA assay was performed. Multiple comparisons using Kruskal–Wallis’s test for IFN-γ CD4^+^ and IFN-γ CD4^+^ CD^8+^ showed that there was a significant increase in vaccination cohort with AZD1222 (*p*<0.0001) or BNT162b2 (*p*<0.0001) within 14 days, as compared with baseline, except that BBIBP was not significantly different. Overall, a T-cell response against SARS-CoV-2 peptides was pronounced in the AZD1222 and BNT162b2 cohort (Figure 4A-B, Supplementary Table S2).

**Figure 4.**
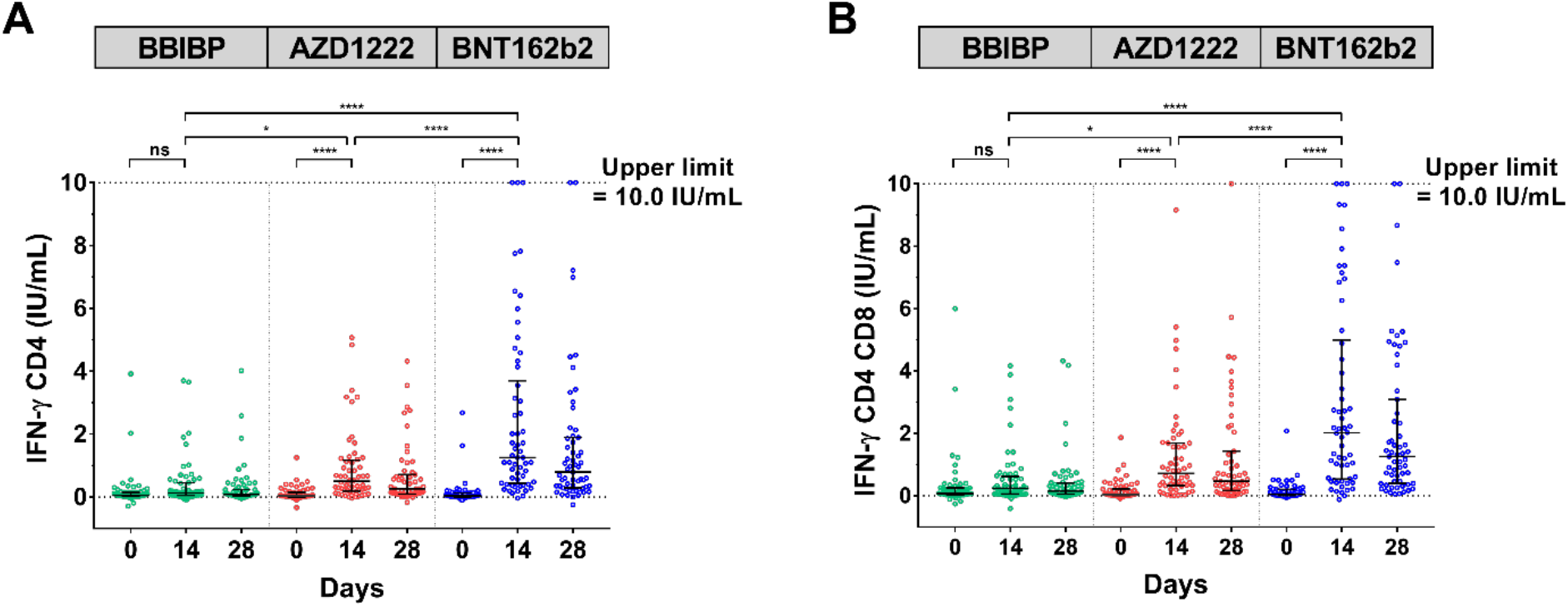
SARS-CoV-2 stimulating IFN-γ assay. The heparinized samples were obtained from participants who received two completed doses of the inactivated vaccine, CoronaVac, followed by the inactivated vaccine, BBIBP (green), the viral-vector vaccine, AZD1222 (red), or the mRNA vaccine, BNT162b2 (blue) at 3–4 months after the first dose, and incubated in QFN blood collecting tube for 21 h. The plasma fraction was evaluated by QFN IFN-γ ELISA. The IFN-γ produced by CD4 specific Ag1 (A); The IFN-γ produced by CD4 and CD8 specific Ag2 (B). Lines represent median (IQR). ns indicates no significant difference; *p*<0.05 (*), 0.0001 (****).

## 4. Discussion

Given the ongoing threat of the SARS-CoV-2 pandemic and its global transmission, vaccines for COVID-19 were urgently developed in early 2021 and authorized for administration to the eligible population worldwide. The driven heterologous COVID-19 vaccines are currently considered its safe and efficiency. This prospective study aimed to evaluate whether the combination of COVID-19 vaccines is safe and required for long-term protection against SARS-CoV-2. The additional booster dose was allowed to be given to the healthy adult population in multiple countries [19–20]. Our data confirmed that mixed and matched vaccines with a complete primary dose of CV followed by BBIBP, AZD1222, or BNT162b2 was safe without serious AEs.

Several prior research studies have focused on the antibody response elicited by the third dose of a similar booster vaccine [21–23]. Recent data have shown that a complete dose of CV could not very efficiently activate an immune response, manifested by a low level of antibody (GMT approximately 100 U/mL at 4 weeks after vaccination) [13, 24–25] and neutralizing activity (approximately 70% WT and nearly 50% variants of concern [VOCs]) [13]. Consistent with our study, the baseline characteristics of their participants after 2–3 months showed lower Ig/IgG anti-RBD and neutralizing activity against SARS-CoV-2 and their variants. Indeed, the third dose with any platform of the COVID-19 vaccine ensured higher immunogenicity. Our cohort study of AZD1222 booster was also similar as previously reported in a real-world study [13]. Additionally, the antibody response after booster with intramuscular BNT162b2 in our current study (3,821 BAU/mL) was similar to previous studies (3,884 BAU/mL) [26]. Previous studies have also found that the mix and match of the platform, namely CV/AZD1222 significantly elicited IgG anti-RBD at 28 days compared to CV/CV [27]. Furthermore, AZD1222/BNT162b synergistically promoted IgG anti-spike at both 28 days [28] and 73 days [29]. In addition, the booster vaccine, especially with AZD1222 and BNT162b2, could develop immune response both IgA anti-spike and IFN-γ stimulation. In accord with this study, the previous report revealed that the group boost BNT162b enhanced immunoglobulin subclass IgG and IgA, and significantly stimulate IFN-γ [29]. Intriguingly, sera IgA anti-spike became dominantly evident in individuals vaccinated with the booster. Previous reports have shown that serum IgA acted as a potent and early SARS-CoV-2–neutralizing agent [30–31]. Nevertheless, we did not observe the titer of vaccine-induced mucosal IgA. This reason for this is supported by the evidence that IgA presents faster than IgG at the mucosal site, and most of the tissue would play an important role as a first-line barrier against infection [31–32]. However, it remains unclear whether AZD1222 and BNT162b2 vaccine induced serum IgA [29,33].

Our study has shown that two doses of inactivate vaccines comprising CV/CV and followed by additional booster with BBIBP at 3-month interval can significantly elicit the IgG anti-N as well as SARS-CoV-2 natural infection. The result can be explained by the fact that the inactivated vaccines—CV and BBIBP—have the SARS-CoV-2 nucleocapsid. While immunization with either AZD1222 or BNT162b2, which does not contain nucleoprotein, did not exhibit any IgG anti-N response. The coronavirus nucleoprotein has been reported to be involved in various aspects of virus replication. A confocal microscopy has shown that the avian infectious bronchitis virus N protein localizes both to the cytoplasmic and nucleolar compartments [34]. Rationally, it has been documented that IgG anti-N is functionally involved in humoral immune response [35], interfering with nucleoprotein localization [34]. Moreover, additional evidence has shown that anti-N sera act as intracellular neutralization required by the cytosolic Fc receptor and E3 ubiquitin ligase TRIM21 [36].

The extracellular neutralization of SARS-CoV-2 and their variants (B.1.1.7, B.1.351, B.1.617.2) were also validated by sVNT. This recent study outlined that vaccinated sera could neutralize WT> B.1.1.7> B.1.617.2> B.1.351 in a similar pattern with three boosters of inactivated vaccine [37], consistent with previous findings in a real-world study [13].

IFN-γ is a potential immunoregulatory protein that facilitates viral clearance brought about by T cells, including T helper type 1 (Th-1) cells, cytotoxic T lymphocytes (CTLs), and natural killer (NK) cells. The current study has shown that T cells produce IFN-γ early, by 14 days, which was notably observed in both the BNT162b and AZD1222 groups, but not the BBIBP group. Previous studies showed that an AZD1222 vaccination generated CD4+-and CD8+-mediated IFN-γ production within 14 days [38]. BNT162b2 vaccination resulted in the detection of IFN-γ CD4^+^ and CD8^+^ cells at 29 days post-boost using the ELISpot assay [39]. Furthermore, evidence has shown that there were supportive T-cell responses in the heterologous AZD1222/BNT162b2 participants [40]. In the present study, the third booster of inactivated BBIBP vaccine stimulated mild CD4+ and CD8+ induction of IFN-γ when compared with the two others described above.

The recent evidence has shown that the neutralizing antibody gradually decrease after two doses of inactivated vaccines, and significantly increase after receiving of the booster dose. A previous study reported that the memory of IFN-γ T-cells against S, N, M, O antigens of SARS-CoV-2 can rapidly awaken after receiving of the third dose of inactivated vaccine [41]. The lately VOC is known as Omicron variant B.1.1.529. This variant is characterized by over 30 changes in spike protein, in particularly 15 changes in RBD resulting in an evasion of host cell recognition and targeted immune response. Many of the changes may potentially increase the transmissibility of the Omicron variant compared to the Delta and Alpha variants [42]. The advantages of the inactivated vaccine in countering the new variant are still under investigation.

A number of caveats need to be mentioned in this study. First, the relatively small sample size limits the statistical power of our results. Therefore, multi-center prospective longitudinal investigations with larger sample sizes should be further investigated to determine whether the data represent more reliability and rare adverse events may be observed, such as thrombosis in AZD1222 and myocarditis in BNT162b2. Second, the combination and prime/boost interval of enrolled participants varied in individual studies. Third, we did not determine live virus focus reduction neutralization tests (FRNTs) in these subjects. However, neutralizing activity against SARS-CoV-2 WT and their variant strains has been examined by an ELISA-based surrogate virus neutralization test. Additionally, there were limitations in quantification of serum IgA as some of the values exceed the upper limit of quantification. Lastly, we did not evaluate the mucosal IgA that reflected the first line barrier against SARS-CoV-2 internalization. Further studies will be needed to overcome these limitations

## 5. Conclusions

The clinical trial of the third booster was a practical design to propose guidelines and build confidence in over 40 countries that administer vast quantities of CoronaVac vaccines. The combination with different platforms as in the present study has yielded immune response against the COVID-19 outbreak with acceptable AEs. Last, the safety and efficacy of heterologous vaccine regimens are guidelines of vaccination practice.

## Data Availability

All data produced in the present work are contained in the manuscript

## Institutional Review Board Statement

The study protocol was approved by the Institutional Review Board (IRB), Faculty of Medicine, Chulalongkorn University (IRB number 546/64)

## Informed Consent Statement

Informed consent was obtained before participant enrollment. The study was conducted according to the Declaration of Helsinki and the principle of Good Clinical Practice Guidelines (ICH-GCP).

## Data Availability Statement

The datasets generated during and/or analyzed during the current study are available from the corresponding author on reasonable request.

## Acknowledgments

We would like to thank all the staff of the Center of Excellence in Clinical Virology and all the participants for helping and supporting in this project. This research was financially supported by Health Systems Research Institute (HSRI), National Research Council of Thailand (NRCT), the Center of Excellence in Clinical Virology, Chulalongkorn University, and King Chulalongkorn Memorial Hospital and partially supported by the Second Century Fund (C2F) to S.K., Chulalongkorn University.

