## supplementary information for "Safety and immunogenicity of the third booster dose with inactivated, viral vector, and mRNA COVID-19 vaccines in fully immunized healthy adults with inactivated vaccine"

**Supplementary Figure and Tables**

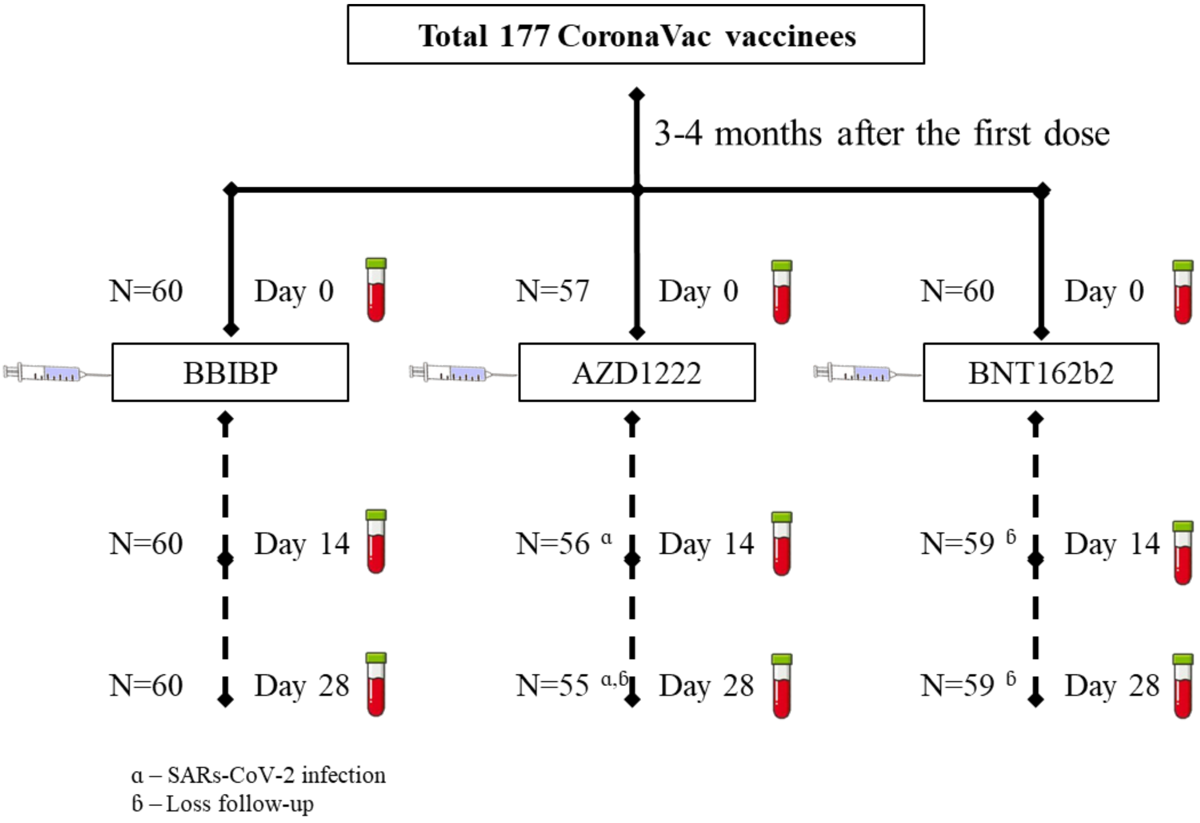


**Supplementary Figure S1.** The participant flow diagram of this clinical study of third booster and blood sampling collection.

| | BBIBP | AZD1222 | *p* value | Result | BBIBP | BNT162b2 | *p* value | Result | AZD1222 | BNT162b2 | *p* value | Result |
|---|---|---|---|---|---|---|---|---|---|---|---|---|
| **n** | | | | | | | | | | | | |
| **Total (%)** | 60 (100.00) | 57 (100.00) | | | 60 (100.00) | 60 (100.00) | | | 57 (100.00) | 60 (100.00) | | |
| **Injection site pain** | 33 (55.00) | 52 (91.23) | < 0.0001 | Yes | 33 (55.00) | 59 (98.33) | < 0.0001 | Yes | 52 (91.23) | 59 (98.33) | 0.1079 | No |
| **Swelling** | 6 (10.00) | 25 (43.86) | < 0.0001 | Yes | 6 (10.00) | 25 (41.67) | 0.0001 | Yes | 25 (43.86) | 25 (41.67) | 0.8531 | No |
| **Redness** | 4 (6.67) | 18 (31.58) | 0.0007 | Yes | 4 (6.67) | 21 (35.00) | 0.0002 | Yes | 18 (31.58) | 21 (35.00) | 0.8446 | No |
| **Fever** | 0 (0.00) | 9 (15.79) | < 0.0001 | Yes | 0 (0.00) | 5 (8.33) | 0.0573 | No | 9 (15.79) | 5 (8.33) | 0.2615 | No |
| **Headache** | 10 (16.67) | 41 (71.93) | < 0.0001 | Yes | 10 (16.67) | 34 (56.67) | < 0.0001 | Yes | 41 (71.93) | 34 (56.67) | 0.1226 | No |
| **Myalgia** | 19 (31.67) | 44 (77.19) | < 0.0001 | Yes | 19 (31.67) | 44 (73.33) | < 0.0001 | Yes | 44 (77.19) | 44 (73.33) | 0.6729 | No |
| **Nausea** | 2 (3.33) | 10 (17.54) | 0.0142 | Yes | 2 (3.33) | 11 (18.33) | 0.0159 | Yes | 10 (17.54) | 11 (18.33) | >0.9999 | No |
| **Vomiting** | 0 (0.00) | 3 (5.26) | 0.1125 | No | 0 (0.00) | 5 (8.33) | 0.0573 | No | 3 (5.26) | 5 (8.33) | 0.7175 | No |
| **Diarrhea** | 5 (8.33) | 16 (28.07) | 0.0074 | Yes | 5 (8.33) | 10 (16.67) | 0.2693 | No | 16 (28.07) | 10 (16.67) | 0.1825 | No |
| **Joint pain** | 3 (5.00) | 23 (40.35) | < 0.0001 | Yes | 3 (5.00) | 13 (21.67) | 0.0136 | Yes | 23 (40.35) | 13 (21.67) | 0.0443 | Yes |

| | | | | | | | | | | | | |
|---|---|---|---|---|---|---|---|---|---|---|---|---|
| **Chilling** | 5 (8.33) | 32 (56.14) | < 0.0001 | Yes | 5 (8.33) | 27 (45.00) | < 0.0001 | Yes | 32 (56.14) | 27 (45.00) | 0.2690 | No |
| **Dizziness** | 7 (11.67) | 21 (36.84) | 0.0021 | Yes | 7 (11.67) | 23 (38.33) | 0.0013 | Yes | 21 (36.84) | 23 (38.33) | >0.9999 | No |

**Supplementary Table S1** Statistic analysis of reactogenicity data of between the booster vaccines.

The Fisher's Exact Test was used to interpret the statistical analysis.

**Supplementary Table S2**. The data values from laboratory testing of participants who received the booster dose with BBIBP, AZD122 or BNT162b2, which referred to graphic info displays.

| | BBIBP | AZD1222 | BNT162b2 |
|---|---|---|---|
| **Humoral responses** | | | |
| **Ig anti-RBD (U/mL)** | | | |
| Day 0 (baseline), n | 60 | 57 | 60 |
| GMT (95% CI) | 35.45 (25.8 - 48.71) | 37.89 (27.39 - 52.41) | 48.57 (36.54 - 64.58) |
| Day 14, n | 60 | 56 | 59 |
| GMT (95% CI) | 1,073 (849.4 - 1,355) | 9,865 (7,990 - 12,182) | 20,787 (18,229 - 23,703) |
| Day 28, n | 60 | 55 | 59 |
| GMT (95% CI) | 839.9 (674.1 - 1,047) | 8,160 (6,635-10,035) | 13,871 (11,993 - 16,043) |
| **IgG anti-RBD (BAU/mL)** | | | |
| Day 0 (baseline), n | 60 | 57 | 60 |
| GMT (95% CI) | 42.76 (33.78 - 54.12) | 41.13 (33.22 - 50.93) | 48.99 (40.77 - 58.88) |
| Day 14, n | 60 | 56 | 59 |
| GMT (95% CI) | 205.5 (167.0 - 253.0) | 1,936 (1,597 - 2,346) | 3,821 (3,306 - 4,416) |
| Day 28, n | 60 | 55 | 59 |
| GMT (95% CI) | 164.1 (133.8 - 201.1) | 1,736 (1,434 - 2,101) | 2,584 (2,250 - 2,966) |
| **IgG anti-N index (S/C)** | | | |
| Day 0 (baseline), n | 60 | 57 | 60 |
| Median (IQR) | 0.265 (0.140 - 1.130) | 0.230 (0.095 - 0.810) | 0.535 (0.170 - 1.078) |
| Day 14, n | 60 | 56 | 59 |
| Median (IQR) | 2.970 (1.715 - 4.745) | 0.205 (0.083 - 0.615) | 0.51 (0.180 - 0.880) |
| Day 28, n | 60 | 55 | 59 |
| Median (IQR) | 2.780 (1.513 - 4.643) | 0.170 (0.070 - 0.400) | 0.51 (0.230 - 0.880) |
| **IgA anti-S1 ratio (S/C)** | | | |
| Day 0 (baseline), n | 20 | 20 | 20 |
| Median (IQR) | 0.685 (0.453 - 0.845) | 0.455 (0.255 - 0.628) | 0.675 (0.453 - 0.845) |
| Day 14, n | 60 | 56 | 59 |
| Median (IQR) | 2.055 (1.073 - 4.383) | 8.515 (5.23 – 9.00) | 9.00 (9.00 - 9.00) |
| Day 28, n | 60 | 55 | 59 |

| | | | |
|---|---|---|---|
| Median (IQR) | 1.300 (0.565 - 2.955) | 7.450 (4.09 – 9.00) | 9.00 (7.12– 9.00) |
| **Neutralization assay sVNT-WT (%inhibition)** | | | |
| Day 0 (baseline), n | 10 | 10 | 10 |
| Median (IQR) | 20.9 (18.33 - 36.95) | 35.15 (19.23 - 46.45) | 40.25 (20.48 - 45.93) |
| Day 14, n | N/D | N/D | N/D |
| Median (IQR) | | | |
| Day 28, n | 30 | 30 | 30 |
| Median (IQR) | 79.75 (62.33 - 89.93) | 97.60 (97.20 - 97.80) | 97.80 (97.68 - 97.90) |
| **Neutralization assay sVNT-Alpha (%inhibition)** | | | |
| Day 0 (baseline), n | 10 | 10 | 10 |
| Median (IQR) | 9.25 (4.50 - 18.53) | 17.30 (17.30 - 26.08) | 19.45 (12.15 - 25.03) |
| Day 14, n | N/D | N/D | N/D |
| Median (IQR) | | | |
| Day 28, n | 30 | 30 | 30 |
| Median (IQR) | 67.20 (51.10 - 81.80) | 96.50 (94.58 - 97.33) | 97.05 (97.05 - 97.53) |
| **Neutralization assay sVNT-Beta (%inhibition)** | | | |
| Day 0 (baseline), n | 10 | 10 | 10 |
| Median (IQR) | -3.80 ((-8.28) - 2.50) | -0.15 ((-19.93) - 10.68) | 1.15 ((-10.95) - 7.50) |
| Day 14, n | N/D | N/D | N/D |
| Median (IQR) | | | |
| Day 28, n | 30 | 30 | 30 |
| Median (IQR) | 50.55 (36.35 - 67.18) | 91.85 (87.80 - 93.98) | 93.30 (90.65 - 94.90) |
| **Neutralization assay sVNT-Delta (%inhibition)** | | | |
| Day 0 (baseline), n | 10 | 10 | 10 |
| Median (IQR) | 24.40 (17.98 - 30.13) | 30.95 (20.70 - 42.65) | 27.40 (20.15 - 40.43) |
| Day 14, n | N/D | N/D | N/D |
| Median (IQR) | | | |

| | | | |
|---|---|---|---|
| Day 28, n | 30 | 30 | 30 |
| Median (IQR) | 72.00 (53.75 - 86.93) | 97.35 (96.45 - 97.73) | 97.60 (97.18 - 97.80) |
| **T-cell responses** <br> **IFN-γ CD4$^+$ (IU/mL)** | | | |
| Day 0 (baseline), n | 60 | 57 | 60 |
| Median (IQR) | 0.050 (0.020 - 0.140) | 0.030 (0.000 - 0.145) | 0.035 (0.000 – 0.128) |
| Day 14, n | 60 | 56 | 58* |
| Median (IQR) | 0.125 (0.053 - 0.453) | 0.500 (0.188 - 1.173) | 1.250 (0.430 - 3.698) |
| Day 28, n | 60 | 55 | 59 |
| Median (IQR) | 0.085 (0.030 - 0.238) | 0.260 (0.090 - 0.710) | 0.790 (0.290 - 1.890) |
| **IFN-γ CD4$^+$ CD8$^+$ (IU/mL)** | | | |
| Day 0 (baseline), n | 60 | 57 | 60 |
| Median (IQR) | 0.075 (0.030 - 0.258) | 0.040 (0.010 - 0.210) | 0.050 (0.010 - 0.208) |
| Day 14, n | 60 | 56 | 58* |
| Median (IQR) | 0.240 (0.060 - 0.615) | 0.720 (0.325 - 1.690) | 2.020 (0.538 - 4.993) |
| Day 28, n | 60 | 55 | 59 |
| Median (IQR) | 0.155 (0.053 - 0.410) | 0.470 (0.170 - 1.430) | 1.260 (0.400 - 3.090) |

N/D – do not determine.
* – one participant's sample is not adequate to evaluate QFN assay because we were unable to obtain the appropriate amount of heparinized blood.
